# Using Deep Learning to Automate Eosinophil Counting in Pediatric Ulcerative Colitis Histopathological Images

**DOI:** 10.1101/2024.04.03.24305251

**Authors:** James Reigle, Oscar Lopez-Nunez, Erik Drysdale, Dua Abuquteish, Xiaoxuan Liu, Juan Putra, Lauren Erdman, Anne M. Griffiths, Surya Prasath, Iram Siddiqui, Jasbir Dhaliwal

**Affiliations:** Division of Gastroenterology, Hepatology and Nutrition, Cincinnati Children’s Hospital Medical Center, Cincinnati, OH, United States; Division of Biomedical Informatics, Cincinnati Children’s Hospital Medical Center, Cincinnati, OH, United States; Department of Biomedical Informatics, University of Cincinnati, Cincinnati, OH, 45229, USA; Department of Pediatrics, University of Cincinnati, College of Medicine, Cincinnati, OH, United States; Division of Pathology and Laboratory Medicine, Cincinnati Children’s Hospital Medical Center, OH, United States; Department of Pathology and Laboratory Medicine, University of Cincinnati College of Medicine, Cincinnati, OH, United States; AI in Medicine for Kids, The Hospital for Sick Children, Toronto, ON, Canada; James M. Anderson Center, Cincinnati Children’s Hospital Medical Center; Department of Pediatric Laboratory Medicine & Pathobiology, Division of Pathology, The Hospital for Sick Children, Toronto, ON, Canada; Department of Pathology, Boston Children’s Hospital, Boston, MA; SickKids IBD Centre, Division of Gastroenterology, Hepatology and Nutrition, The Hospital for Sick Children, Department of Pediatrics, University of Toronto, Toronto, Canada

**Keywords:** Deep learning, cell counting, histopathology imaging, eosinophilic cells, ulcer, inflammatory bowel disease, ulcerative colitis

## Abstract

**Background:** Accurate identification of inflammatory cells from mucosal histopathology images is important in diagnosing ulcerative colitis. The identification of eosinophils in the colonic mucosa has been associated with disease course. Cell counting is not only time-consuming but can also be subjective to human biases. In this study we developed an automatic eosinophilic cell counting tool from mucosal histopathology images, using deep learning.

**Method:** Four pediatric IBD pathologists from two North American pediatric hospitals annotated 530 crops from 143 standard-of-care hematoxylin and eosin (H & E) rectal mucosal biopsies. A 305/75 split was used for training/validation to develop and optimize a U-Net based deep learning model, and 150 crops were used as a test set. The U-Net model was then compared to SAU-Net, a state-of-the-art U-Net variant. We undertook post-processing steps, namely, (1) the pixel-level probability threshold, (2) the minimum number of clustered pixels to designate a cell, and (3) the connectivity. Experiments were run to optimize model parameters using AUROC and cross-entropy loss as the performance metrics.

**Results:** The F1-score was 0.86 (95%CI:0.79-0.91) (Precision: 0.77 (95%CI:0.70-0.83), Recall: 0.96 (95%CI:0.93-0.99)) to identify eosinophils as compared to an F1-score of 0.2 (95%CI:0.13-0.26) for SAU-Net (Precision: 0.38 (95%CI:0.31-0.46), Recall: 0.13 (95%CI:0.08-0.19)). The inter-rater reliability was 0.96 (95%CI:0.93-0.97). The correlation between two pathologists and the algorithm was 0.89 (95%CI:0.82-0.94) and 0.88 (95%CI:0.80-0.94) respectively.

**Conclusion:** Our results indicate that deep learning-based automated eosinophilic cell counting can obtain a robust level of accuracy with a high degree of concordance with manual expert annotations.

## 1. Introduction

Eosinophils are pleiotropic multifunctional leukocytes involved in initiating and propagating diverse inflammatory responses and modulating innate and adaptive immunity [1]. Recent studies have shown a role for eosinophils in predicting disease course and response to therapy in ulcerative colitis (UC), a chronic disease of the gastrointestinal tract [2, 3]. The quantification of the number and location of eosinophils in UC from histopathological imaging data could be instrumental in both assessing disease progression and determining the most effective treatment.

The current approach by pathologists is to identify and quantify the peak eosinophilic count (a focused area in the image) rather than counting the entire number of cells in the whole slide image. Manual cell counting is time-consuming and subject to inter- and intra-observer reproducibility, which then raises the question as to whether this can be overcome by advanced computational approaches [4–7].

Cell counting algorithms have existed for some time as reviewed by Xing et al[8]. Machine learning algorithms for cell counting include Distance Transform [9], morphology operation [10], H-Minima [10] and H-Maxima Transform [11], Laplacian of Gaussian [12], Maximally Stable Extremal Region [13], Hough Transform [14, 15], Radial Symmetry-Based Voting [16, 17], and Supervised Learning (Support Vector Machine [18, 19], Random Forest [20], and Deep Learning [21–23]). Our method builds on the success of deep learning in a variety of image processing tasks. Deep learning (DL) models are widely used in solving problems in the biomedical imaging domain [22, 24] including image processing tasks such as segmentation and classification [25]. In this work, cell segmentation has been extended to apply to colonic mucosal tissue samples, stained for hematoxylin and eosin (H & E). Several DL approaches for cell segmentation/counting exist in the literature [8, 26], and the field is an area of active research interest. While a variety of adaptations have recently been presented in the DL community, the U-Net architecture remains the most widely used deep learning approach for cell segmentation [4].

The U-Net model is an encoder-decoder deep-learning method in which the dimensions of the input and output of the model are identical. U-Net provides an efficient and accurate method to segment (outline) cells at various scales [27]. Histopathology slides represent a unique challenge for DL models for several reasons [28]. First and foremost, the images are very large, typically in the gigapixel range which necessitates breaking the image into smaller images or “patches” to permit analysis with less computational cost. Second, the context of cells at low and high magnifications are often needed, such that images need to be examined at multiple scales [6].

In this work, a deep learning approach has been applied to a multi-center dataset to count eosinophils in histopathology images in the context of ulcerative colitis. Our goal is to develop an end-to-end computational pipeline to accurately localize and count the number of eosinophils within the lamina propria from rectal mucosal whole slides images (WSIs). For this purpose, we employed the U-Net architecture [4] to derive an end-to-end segmentation and cell counting model. We compared our results to SAU-Net [29], a state-of-the-art cell counting algorithm. This work utilizes histopathological images from two institutions to train, validate, and test our pipeline, with expert pathologists from two different institutions providing ground truth annotations.

The remaining paper is organized as follows: In Section 2, the pipeline is delineated in a step-by-step manner. This involved analysis at both the patch-level and slide-level scales. Section 3 presents the application of our end-to-end eosinophil counting approach to a multi-center IBD dataset. Finally, in Section 4, the results of the highlighted study are examined and discussed.

## 2. Methods

The goal of this work is to develop an efficient tool to count and localize eosinophils in histopathology images which could feasibly be deployed for clinical use. As the computational size of the slides prevents training on the whole slide images (WSIs), the images were cropped into patches (typically 500 x 500 pixels) and the DL model was trained and evaluated on these patches. The whole slide images (WSI) were analyzed by tiling slides into a non-overlapping, contiguous grid and aggregating counts and locations of eosinophils in each individual tile, see Section 2.9. The proposed patch-level eosinophilic cell counting pipeline is comprised of six steps with further extension to the slide-level extension, building upon the patch-level cell counting results. The algorithm was trained on a standard PC with an Intel i7 CPU and 64GB of RAM. It should be noted, however, that the pipeline is capable of running in parallel on systems with graphical processing units (GPUs). The entire pipeline was written in Python 3.7 and we utilized PyTorch 1.8.0 for the model backbone [30]. This study was approved by the Institutional Review Boards at each of the participating sites, Cincinnati Children’s Hospital Medical Center, Cincinnati, Ohio and the Hospital for Sick Children, Toronto, Ontario.

**Figure 1:**
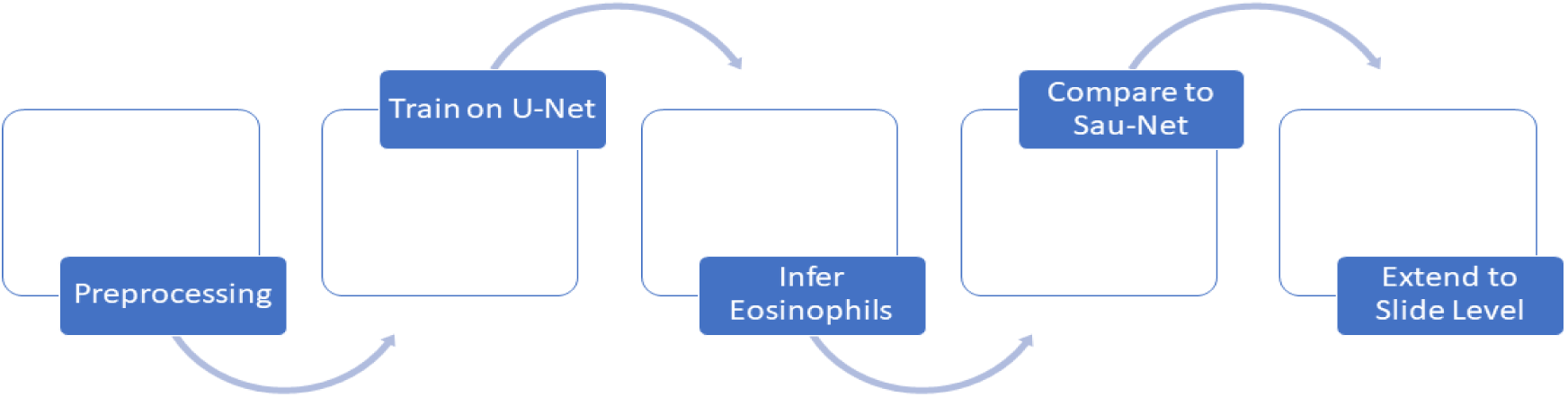
Methodological Pipeline.

### 2.1 Sample Collection and Processing

We used data from two North American pediatric centers, Cincinnati Children’s Hospital Medical Center (Site One) and the Hospital for Sick Children in Toronto, Ontario (Site Two) for model training. For both sites, standard 4-5µm slides of rectal mucosal biopsies were stained with hematoxylin and eosin (H & E, Roche, HE600) [31], then scanned at 20x (Aperio AT2) for digital analysis. The original SVS format pathology slides were converted to PNG without compression for running the subsequent image analysis.

### 2.2 Preprocessing and Patch Generation and Selection

Given the variability across WSIs collected from different centers, we undertook various preprocessing steps using the same method across centers [32]. The steps included removal of excess whitespace wherein the tissue information is sparse, followed by stain normalization, and then data augmentation. WSIs in PNG format were read in greyscale using the OpenCV python library (cv2) [33]. A two-stage Gaussian filter with 4% of the pixels buffered was then used to smooth the image (OpenCV, Numpy) [33, 34]. The midpoint between the minimum and maximum pixels of the image was then used as a threshold to exclude non-informative pixels. 500×500 pixel patches were generated from the smoothed image and used for model development. Only tiles with a brightness threshold greater than 0.95 and whitespace less than 0.88 were selected as model input.

**Figure 2:**
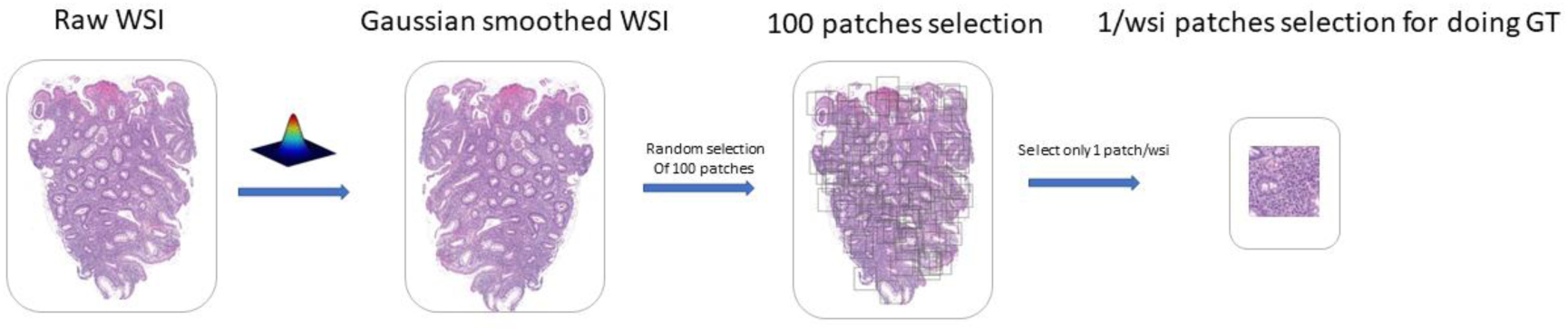
Preprocessing and Patch Generation and Selection from WSIs. We apply a Gaussian filter and apply tiling to obtain non-overlapping patches (500 x 500). For ground-truth generation of eosinophils we randomly selected 100 patches containing tissue across the slides and those were annotated by an IBD pathologist. For DL model training we selected all the patches that had tissue content.

### 2.3 Deep Learning Training and Expert GI Pathologists Annotations

Following pre-processing, patch generation and selection, the next step in the pipeline was to construct the training, validation, and test sets. The model was trained including both Site one and Site two slides with an 80/20 training/validation split (305/75). The training/validation split was chosen based on established norms in machine learning and to weigh in favor of model training given the relatively small number of annotated patches. 150 patches were utilized for the test set, which included 54 patches from site one and site two and 96 patches from a single patient from site one annotated by a single pathologist.

Whole slide images were then divided into 500 x 500-pixel patches. In total, we had 530 annotated patches, 88 from site one and 292 from site two, and a single site one whole slide image with 96 patches, and then 54 test patches that were annotated by two expert pathologists. Table 1 shows the number of patients, for training, validation, and test splits across the sites.

**Table 1:** Number of Patients in our Datasets and Train/Validation/Test Sets Across the Two Sites Used in Testing DL Models.

|  | **Total** | **Training** | **Validation** | **Test** |
| --- | --- | --- | --- | --- |
| **No of Patients** | **143** | **97** | **64** | **51\*** |
| **Site One** | 17.4% | 22.7% | 23.4% | 15.7% |
| **Site Two** | 82.6% | 77.3% | 76.6% | 84.3% |
| **No of Patches** | **530** | **305** | **75** | **150** |
| **Site One** | 36.0% | 23.3% | 22.7% | 53.9% |
| **Site Two** | 64.0% | 76.7% | 77.3% | 46.1% |
| *For test set included one patient from site one, whereby 96 patches were annotated |  |  |  |  |

Following patch generation, the next step in the pipeline was to establish the ground truth (GT) of the training images. One hundred informative 500 x 500-pixel patches for each WSI were randomly sampled with the mean number of patches per patient being 3.2 with a median and interquartile range of 3 (2,4). Four expert pathologists were utilized to annotate the number and location of eosinophils in the training and validation images. For the test set, two pathologists were employed to annotate the same set of patches to provide inter-annotator variability. The patches were annotated in QuPath [35] and the number and locations of each eosinophil were exported from the software.

**Figure 3:**
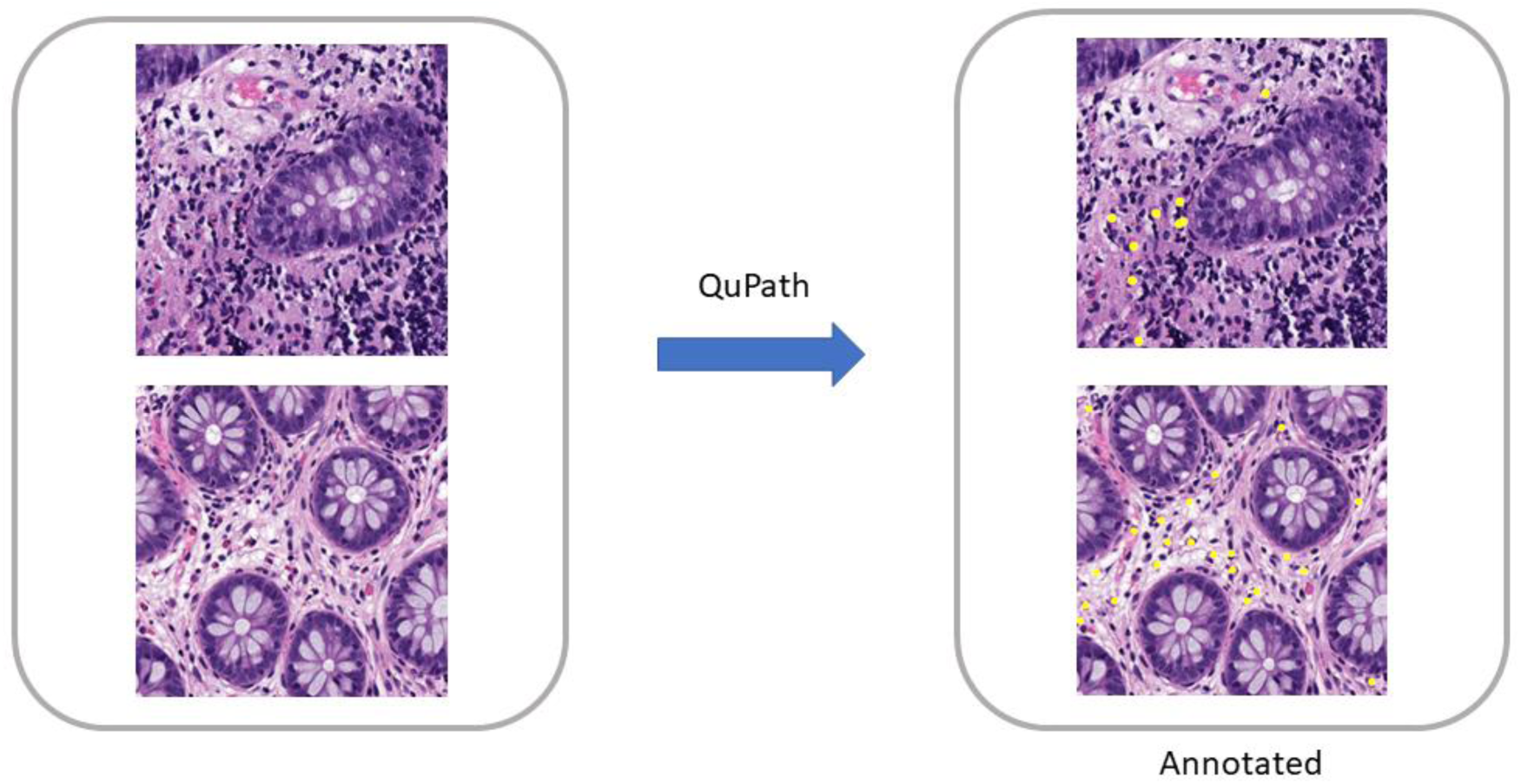
Example annotations on two representative samples of patches. Eosinophil annotations are shown in yellow. Annotated patches and corresponding ground-truth eosinophils were used for training our DL models.

### 2.4 Model Architecture for Cell Counting

A detailed description of the U-Net model used in the pipeline is described in Figure 4. The U-Net architecture has five convolutional layers with the number of channels doubling at every layer. A distinctive feature of the U-Net model is its symmetric architecture, which includes a decoder path with transpose convolutions that restore the dimensions of the output layers to match the original input image size. As a result, the input and output image sizes are the same. Furthermore, U-Net uses skip-connections by appending encoder output to decoder input at every level. The raw U-Net output probabilities were computed using the pixel-wise logistic loss and a post-training threshold was applied to indicate the presence or absence of eosinophils at inference. Parameters were systematically optimized including the learning rate (r), number of epochs, batch numbers, and initial number of U-Net parameters (p) using grid search.

**Figure 4:**
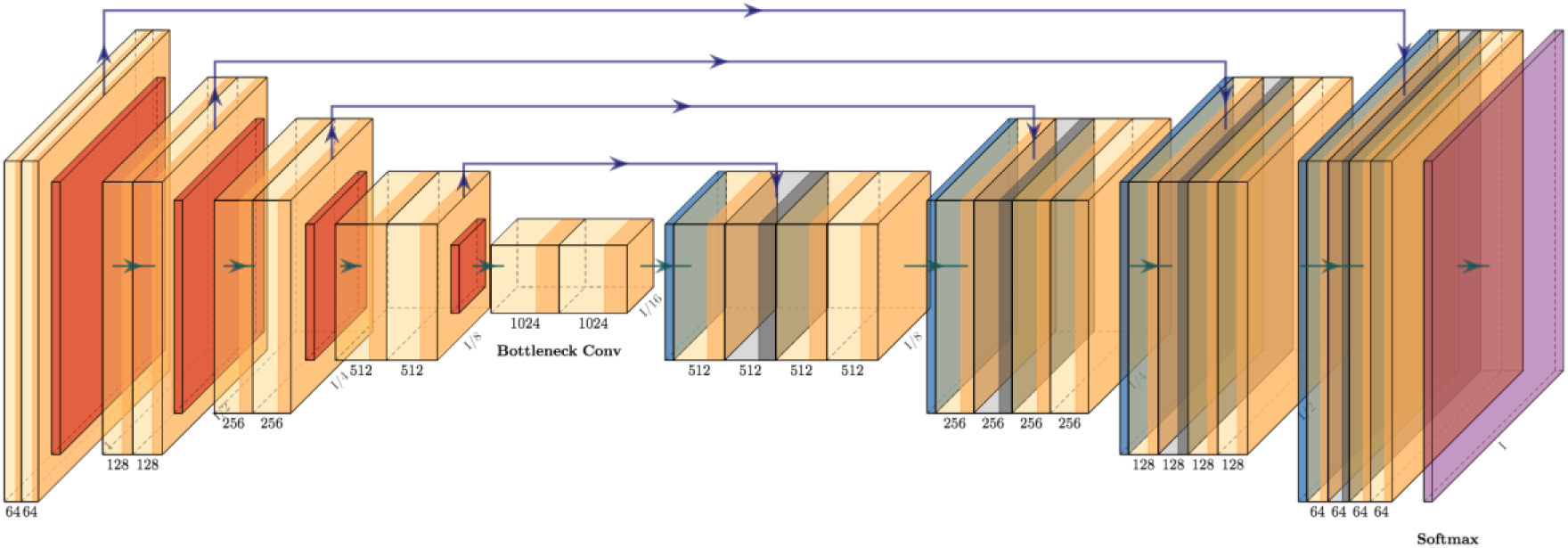
U-Net Deep Learning Architecture [24].

**Figure 5:**
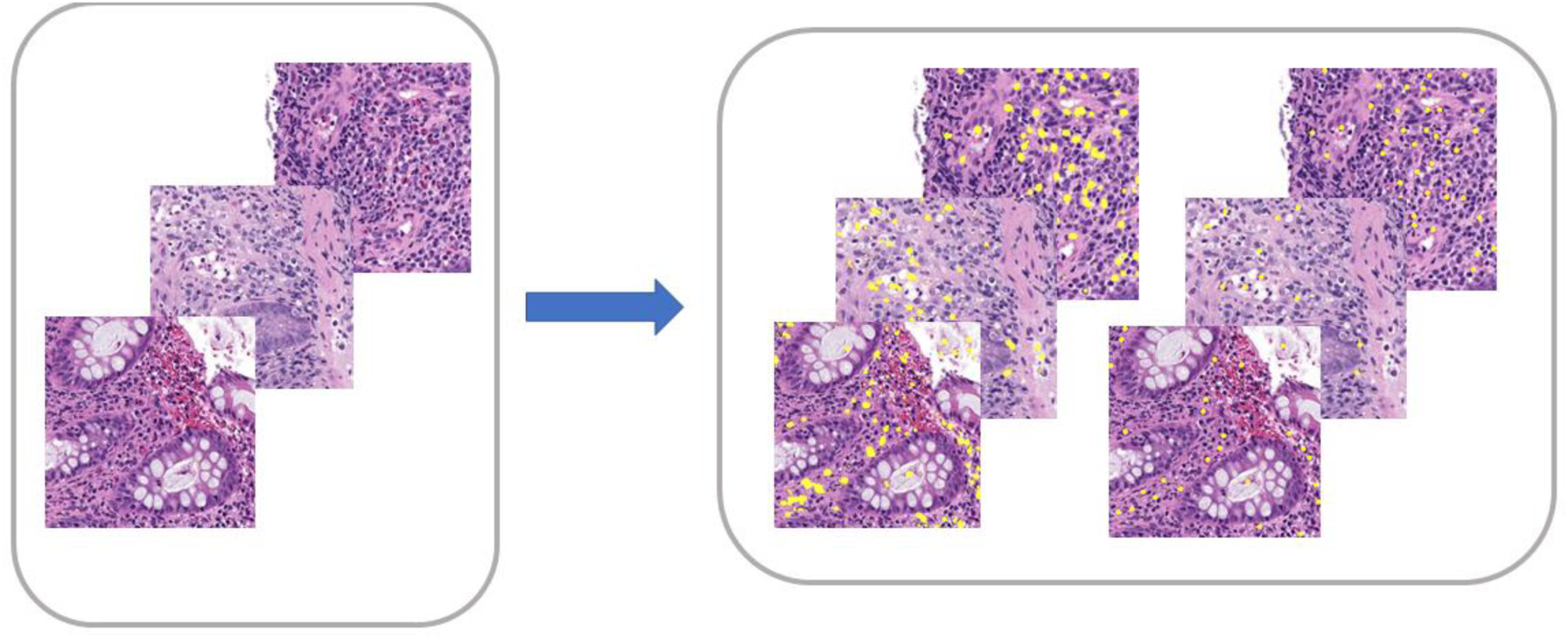
Inference on Test Set Patches Can be Performed for Comparison to Pathologist Annotations.

We briefly describe the U-Net architecture and [4] for more details. The energy function for the network is given by the following equation:

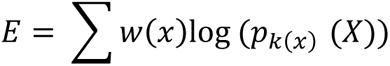

where p_k_ is the pixel-wise SoftMax function applied over the final feature map.

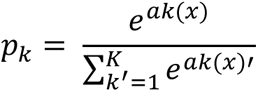

and a_k_(x) denotes the activation in channel k. The U-Net model outputs probabilities of the classes in [0, 1] range and threshold to obtain discrete classes.

### 2.5 Post Processing

In experiments we observed that the application of U-Net model resulted in a large false positive rates for eosinophils, hence a clustering-based post-processing technique was adpated from a previously described approach to differentiate the true positives from the false positives[36]. Clustering was used to group the pixels into related objects. This clustering step can utilize one of two connectivity frameworks, namely 1-connectivity and 2-connectivity. 1-connectivity is a structure consisting of observing the left, right, top, and bottom pixels in relation to a representative pixel. 2-connectivity, on the other hand, observes the corner pixels in addition to the left, right, top, and bottom pixels. Once clusters are determined based on the chosen connectivity framework, a second threshold is then applied that represents the minimum number of grouped pixels to qualify as a cell (eosinophil). As per our extensive experiments, the optimal settings for the connectivity were determined to be 1-connectivity (see Section 3.15) hence that was used in all following analyses. Three parameters optimized were, namely, (1) the pixel-level probability threshold, (2) the minimum number of clustered pixels to designate a cell, and (3) the connectivity.

### 2.6 Inference

The final step of the patch-level pipeline was to use the trained model to predict the number of eosinophils in a separate test set of patches. The pipeline can generate the number and locations of eosinophils in the test set patches.

### 2.7 Evaluation Metrics

Evaluation metrics utilized to optimize the parameters were the area under the receiver operator characteristic curve (AUROC) and the cross entropy loss (CE). Metrics used for model evaluation included pixel-wise AUROC, precision (TP / TP + FP), recall (TP / TP + FN), and F1 (2*Precision*Recall / Precision + Recall). Patch-level results in section 3.2 report the AUROC for the training, validation, and test sets. In addition, to compare the inter-rater variability observed between annotators, the non-parametric Spearman correlation was chosen as the evaluation metric.

### 2.8 Comparison to State-of-the-Art U-Net Variant

To assess the robustness of the pipeline, another state-of-the-art U-Net variant (SAU-Net) was used for training and inference on the same dataset. SAU-Net [29] applies a self-attention module to a U-Net architecture. The model was trained on the same set of patches and inference was performed on the same test set patches. As the output for SAU-Net was the raw cell counts per patch, the evaluation metric we chose to compute for comparison was the mean absolute error.

### 2.9 Extension to Slide Level

One of the central challenges in analyzing histopathology slides is the computational size of the digitized images. Several approaches have been used to address this issue [37–39]. In this pipeline whole slide images were divided into random 500 x 500 pixel patches. These patches were then used as input to train the model. In order to visualize counts across the entire slide an extension to this methodology was employed. Specifically, each slide was tiled into contiguous, non-overlapping 500 x 500 units and the patch-level trained DL model was used to infer eosinophil counts and obtain local coordinates of the eosinophils. To create a unified visualization of the eosinophils overlaid on the original images, the eosinophil locations were transformed to global, slide-level coordinates and overlaid on the original image. Figure 6 shows an example WSI along with our tile selection procedure. We applied two pre-processing steps to remove non-informative tiles. The first step involved removing tiles with a brightness threshold greater than 0.95 (red-colored), followed by a whitespace removal step for tiles with values lower than 0.88 (green-colored). As shown in Figure 6 (middle column) the red tiles were dropped since they contain primarily the white spaces with little or no tissue content, and green tiles constitute edge of the tissue which increased the false-positive eosinophil counts compared to gold-standard pathologists’ annotations as shown in Section 3.4. Finally, the blue tiles are used to show the DL based eosinophils at the WSI level.

**Figure 6:**
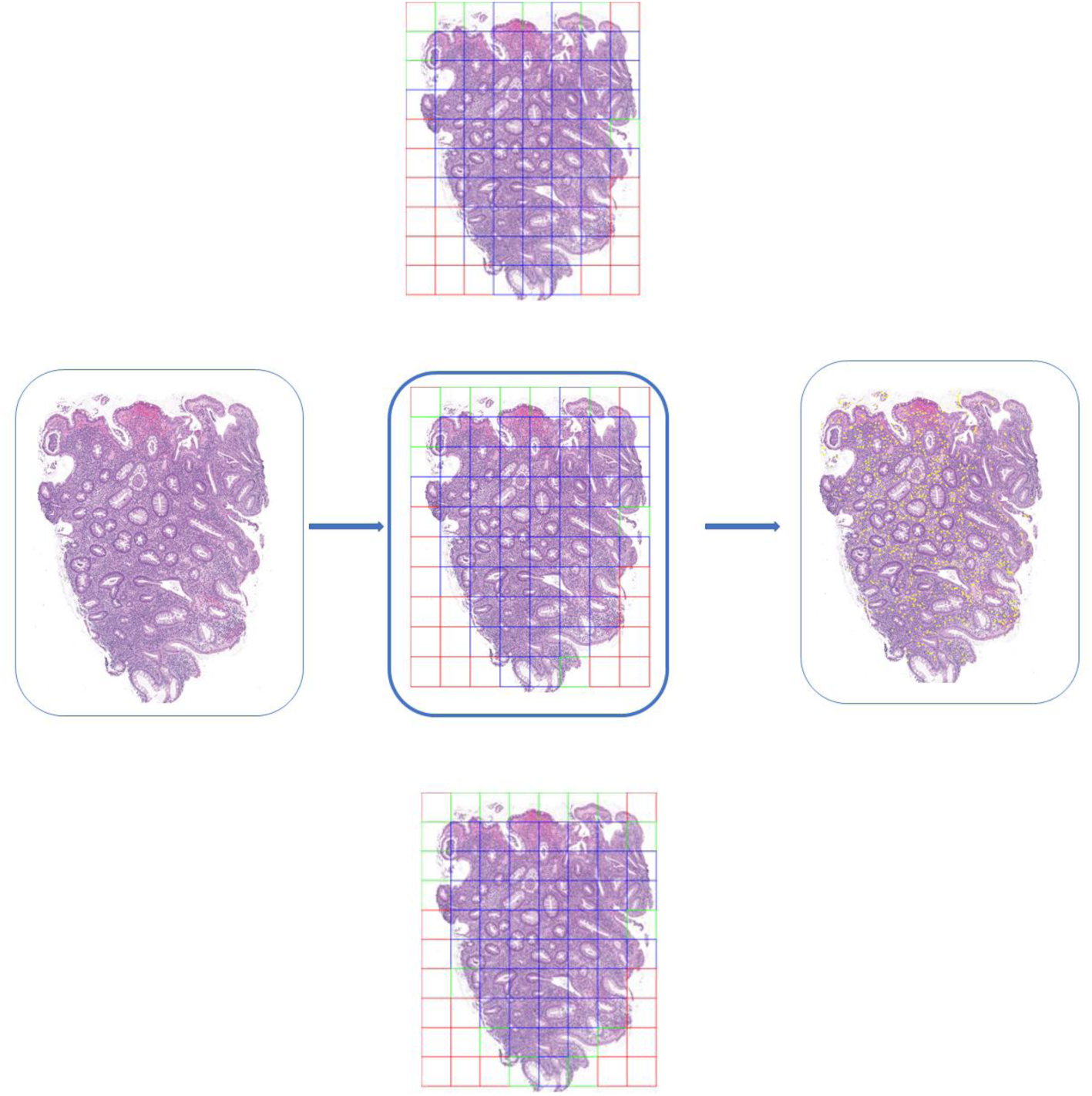
Extension to Slide-Level Inference and Visualization. We keep only tiles which have a brightness threshold greater than 0.95 and then exclude those tiles that have greater than 0.88 whitespace. The middle column shows included tiles for various whitespace removal thresholds. The middle column (top to bottom) shows tiles for Input WSI 0.85, 0.88, 0.90 respectively. Red tiles are dropped based on initial pre-processing, green tiles are excluded after whitespace thresholding, and blue tiles are included in analysis.

## 3. Experiments

### 3.1 Patient Characteristics

Site one data consisted of 25 patients, the median age was 15 years, IQR (13,17), and 44% were male, 100% were white. The UC clinical severity was measured using the physician global assessment score: Quiescent (14.7%), Mild (17.6%), Moderate (11.8%), and Severe (55.9%) (Supplemental Table 1). Site two data consisted of 118 patients, the median age was 13 years IQR (9, 19), 42% were male, 42% Caucasian, and 22% South Asian. The Physician Global Assessment score for site two patients was: Mild (11.9%), Moderate (25.1%), and Severe (38.1%).

### 3.2 Results from Parameter Optimization Experiments

Figure 7A shows the plots of the area under the receiver operator characteristic curve (AUROC) and the cross entropy loss (CE) (scaled against the worst values) loss for parameter optimization. We noticed that when we reduced the learning rate from 0.002 to 0.001, the AUROC dropped from 0.96 (95%CI:0.94-0.98) to 0.94 (95%CI:0.91-0.97) (1.4%) as shown in Figure 7A. Furthermore, when we further reduced the learning parameter to 0.0005, the AUROC dropped significantly to 0.89 (95%CI:0.85-0.93) (7.3%). Batch 6 was chosen as it had the best performance, balancing both AUROC and CE.

**Figure 7:**
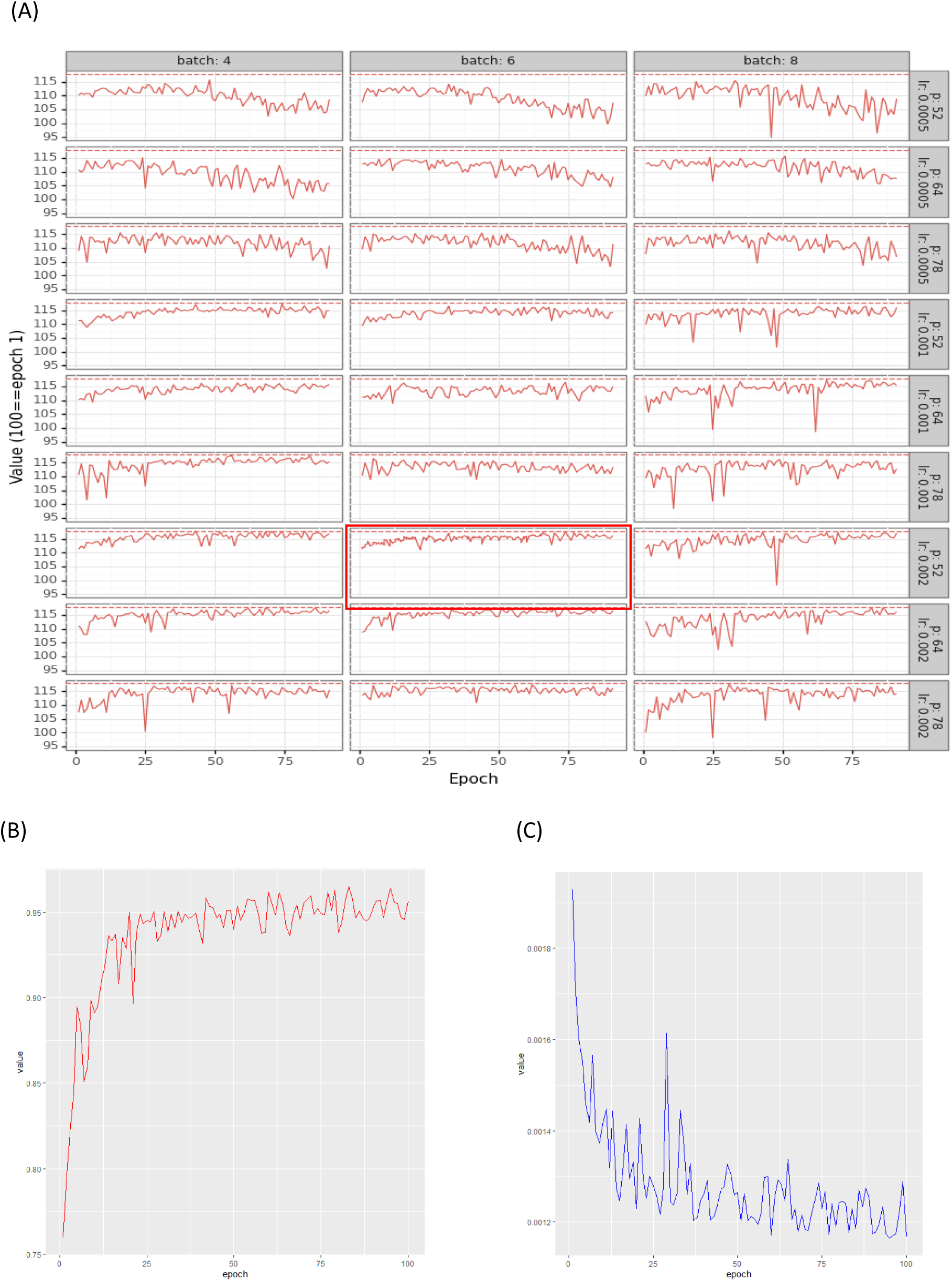
Parameter Sweep to Obtain Optimal Parameters (Panel A) where we show epoch versus accuracy for different batch sizes (4, 6, 8), learning rates (r), and the initial number of UNet parameters (p). Area under the receiver operator curve (AUROC) (Panel B) and Cross Entropy (CE) Loss (Panel C) Curves for Optimal Hyperparameters vs. Number of Training Epochs

**Figure 8:**
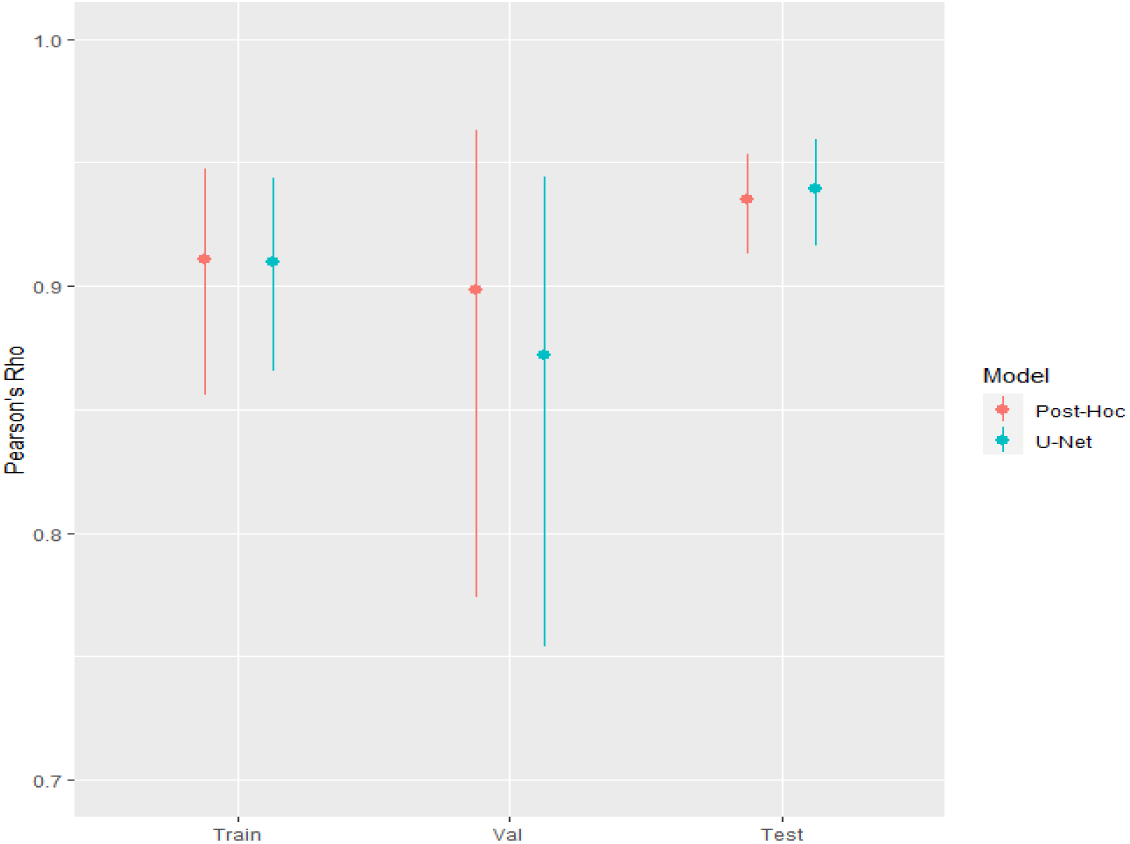
Pearson’s Rho for Comparison of Aggregated Predicted versus Actual Eosinophilic Counts for Training, Validation and Test Sets for Standalone UNet and UNet with Post Processing

Following hyperparameter optimization in our pipeline, the learning rate was set at 0.002, the number of epochs for model training was set at 83, and the batch number was set at 6; while the initial number of U-Net parameters (p) was fixed at 64. The post processing parameters, namely, the pixel-level probability threshold, the number of pixels to designate a cell, and the connectivity, were optimized to be 5.1927 x 10^-3^, 120, and 1, respectively. Figure 7 (B, C) shows AUROC and CE curves for the optimal parameters. Notice that the plateaus at approximately 0.95 (95%CI:0.93-0.97) in the optimized loss curve (Figure 7b) and the cross-entropy loss stabilizes to approximately 0.001 (Figure 7C).

### 3.3 Patch Level Results

#### 3.3.1 Eosinophilic Pipeline Exhibits Robust Results

The eosinophilic-counting pipeline showed robust results with AUROC values of 0.96 (95%CI:0.95-0.96), 0.95 (95%CI:0.94-0.96), and 0.94 (95%CI:0.94-0.95) training, validation, and test, respectively.

The precision and recall of the test set were 0.77 (95%CI:0.70-0.83) and 0.96 (95%CI:0.93-0.99), respectively, yielding an F1 score of 0.86 (95%CI:0.79-0.91).

Figure 9 shows visualizations of eosinophils overlaid on the original patch. Three representative patches were chosen. The first patch (row 1) is an example in which the predicted number (columns, 4 and 5) and locations of eosinophils matched the annotators (columns 2 and 3). Row 2, in turn, portrays a moderate case. Row 3 exhibits an example in which the algorithmic predictions did not closely match the ground truth (Annotators 1 and 2).

**Figure 9:**
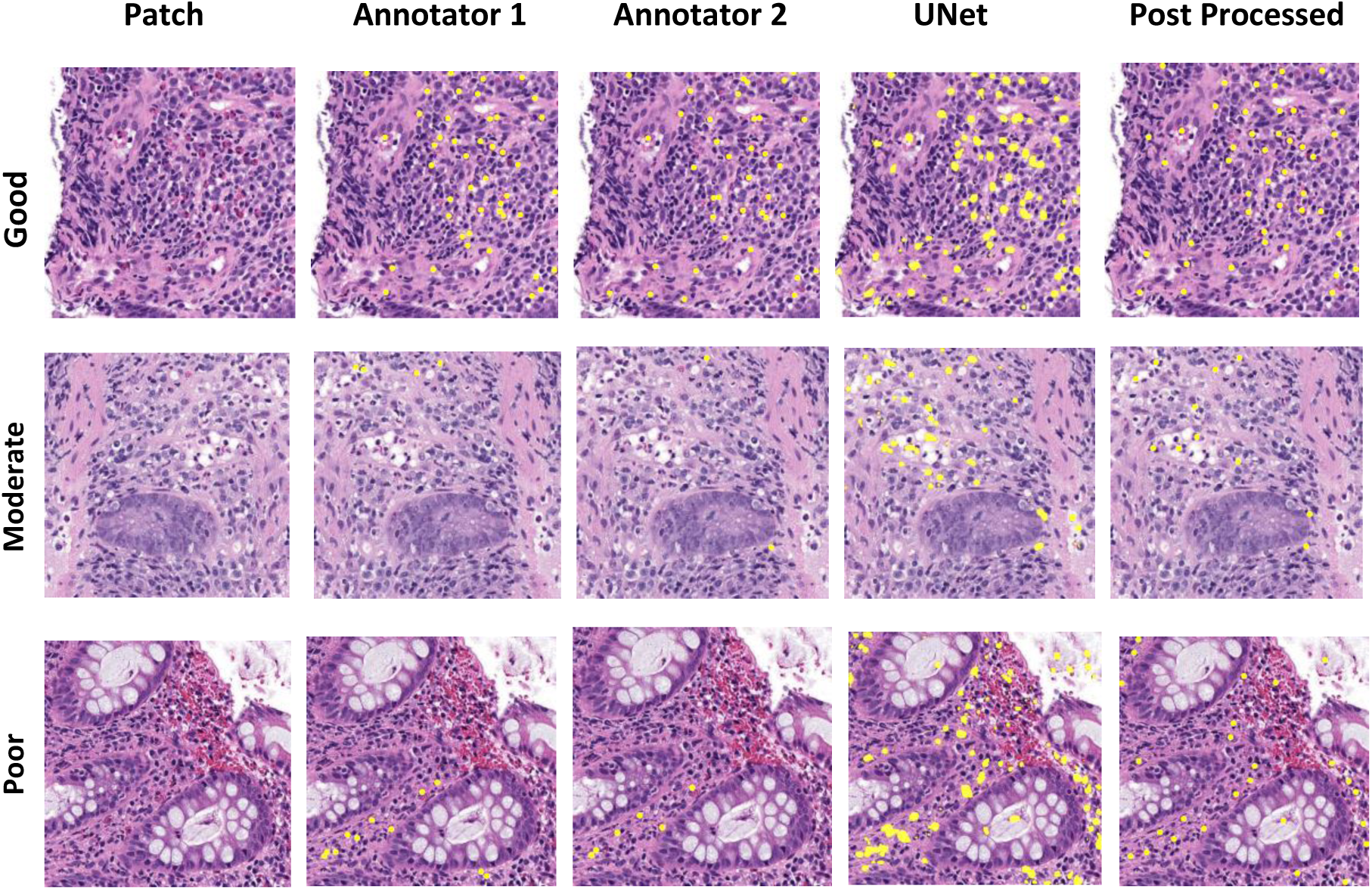
Figure 9 shows visualizations of eosinophils overlaid on the original patch. Three representative patches were chosen. Counts and Visualization of Example Patch (1_st_ column), Annotator 1 (2nd column), Annotator 2 ( 3_rd_ column), U-Net without Post Processing (4_th_ column), and U-Net with Post Processing (5_th_ column) for an example patch. The rows represent (1) good, (2) moderate, and (3) poor examples comparing annotator counts/location to the algorithm. The first patch (row 1) is an example in which the predicted number (columns, 4 and 5) and locations of eosinophils matched the annotators (columns 2 and 3). Row 2, in turn, portrays a moderate case. And row 3 exhibits an example in which the algorithmic predictions did not closely match the ground truth for UNet with or without post processing (Annotators 1 and 2).

#### 3.3.2 Annotators Demonstrate Consistent Results

To accurately assess the performance of the pipeline and the variance between annotators, the correlation between each annotator and the algorithm and between two annotators (OLN, DA) was computed as shown in Figure 10. Annotator 1 showed a 0.88 (95%CI:0.83-0.93) and 0.87 (95%CI:0.82-0.92) correlation coefficient with the U-Net and U-Net plus post-hoc pipelines, respectively. Similarly, Annotator 2 demonstrated a correlation coefficient of 0.89 (95%CI:0.83-0.94) and 0.87 (95%CI:0.82-0.92) between U-Net and U-Net plus post-hoc, respectively. It is noteworthy that the comparison of the two annotators yielded a robust 0.96 correlation, which suggests competence with the ground truth with which the pipeline has been compared.

**Figure 10:**
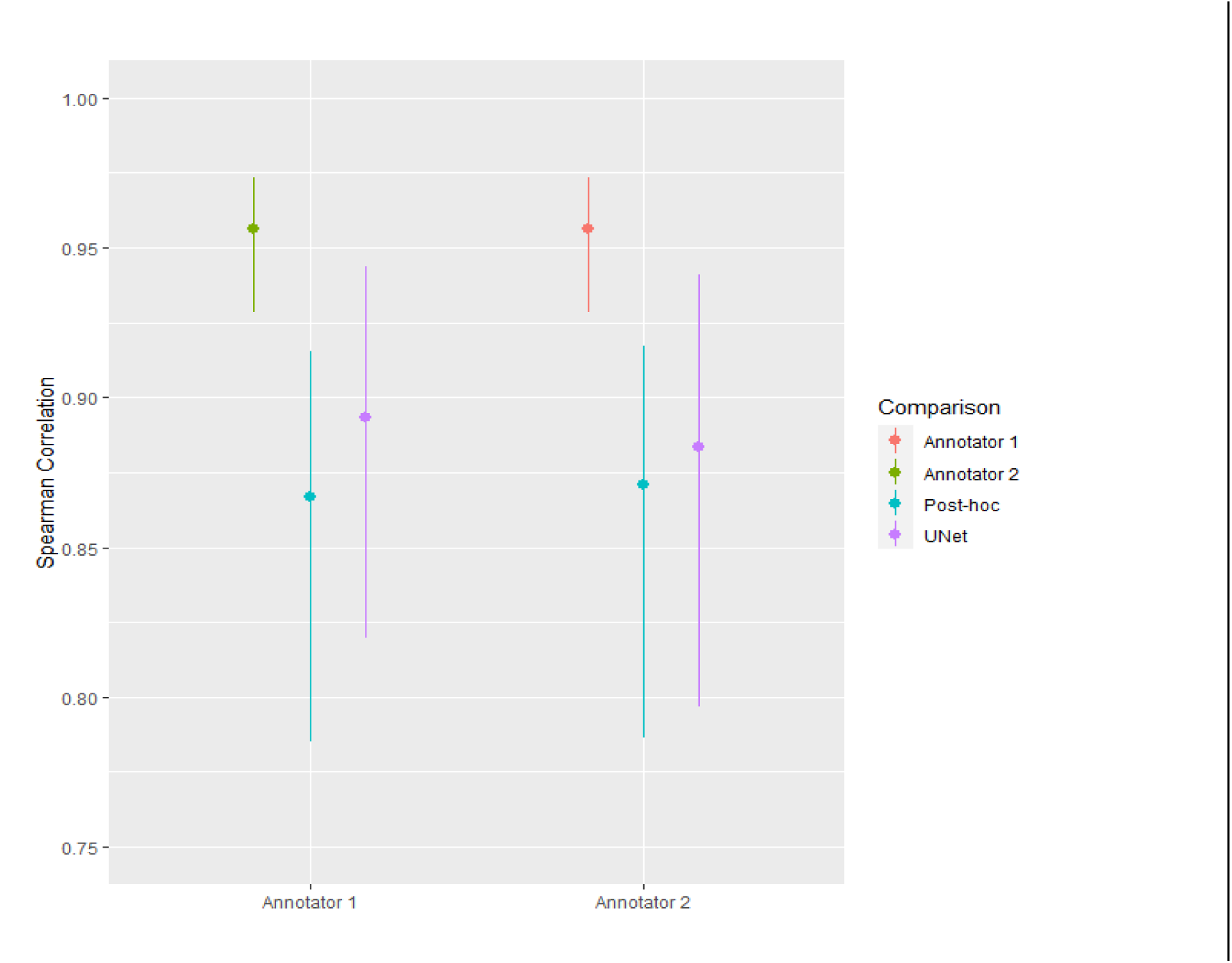
Spearman Correlation of Each Annotator in Relation to Standalone UNet, UNet with Post Processing, and the other Annotator.

#### 3.3.3 Comparison of Patch Level Results to State-of-the-Art U-Net Variant

The AUROC for SAU-Net for the test set was computed to be 0.50 (95%CI:0.42-0.58). The mean absolute error of the pipeline was 3.3 (95%CI:2.6-4.0) as compared to 10.7 (95%CI:9.0-12.4) for SAU-Net. In addition, the precision, recall, and F1 score of the test set yielded values of 0.38 (95%CI:0.31-0.46), 0.13 (95%CI:0.08-0.19), and 0.2 (95%CI:0.13-0.26). These data suggest that in contrast to our pipeline, the SAU-Net algorithm tends to undercount the number of eosinophils, as evidenced by the low number of false positives and high number of false negatives. In addition, a Bland-Altman analysis was performed which suggested two items, (1) our pipeline consistently overcounts cells in relation to SAU-Net, and (2) there was an inverse correlation between the size of the discrepancy between the two methods and the average measurements (Figure 11). In the Bland-Altman plot, the average difference line lying below zero suggests that on average the SAU-Net counts are smaller than our pipeline. Furthermore, the inversely linear relationship observed between the difference between counts and the average of the measurement suggests that our pipeline overcounts in relation to SAU-Net proportionally as the average counts between the two methods increases.

**Figure 11:**
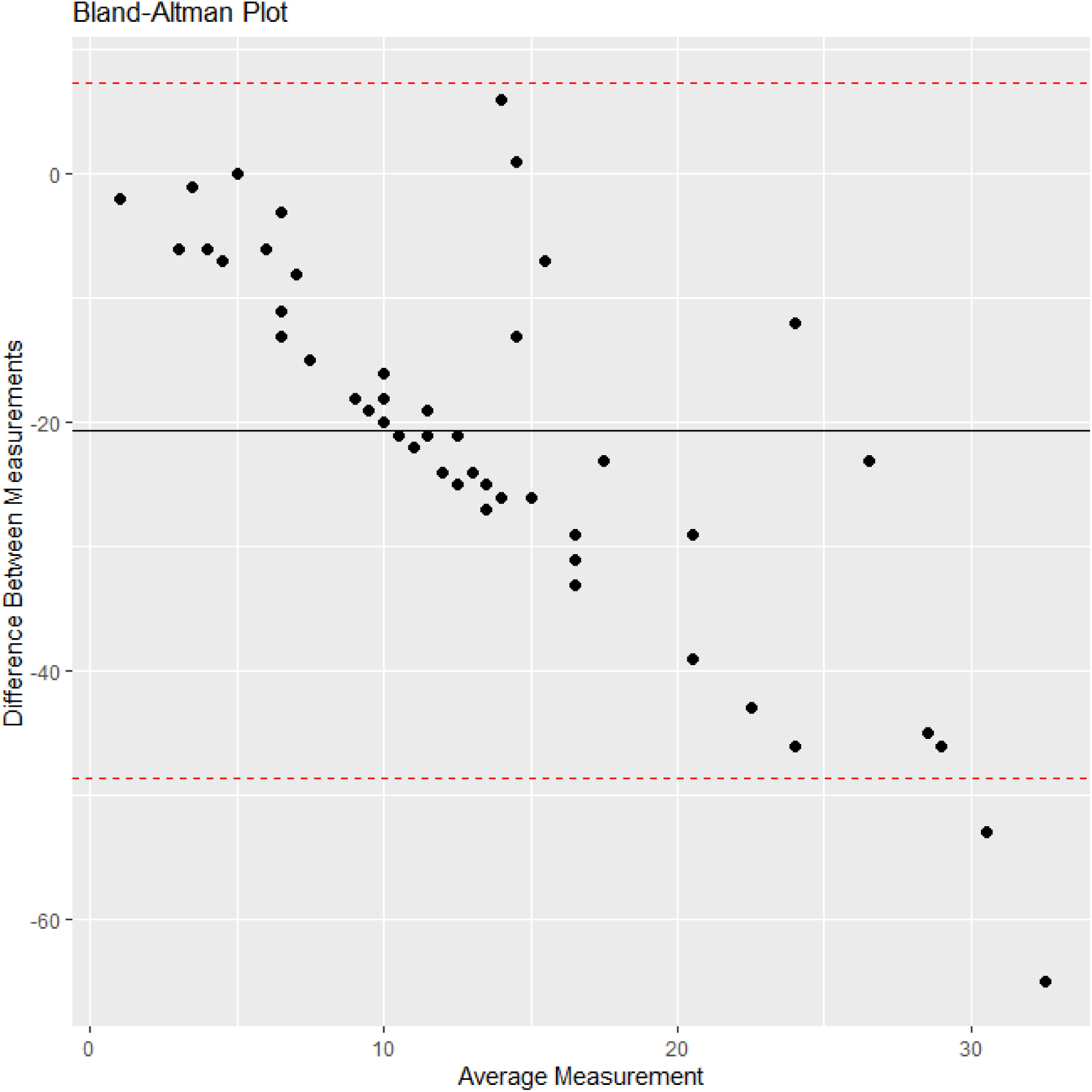
Bland-Altman plot of comparison of UNet with Post Processing Versus SAU-Net for the test set (w/o CCHMC 96). The x-axis portrays the average measurement of the two instruments and the y-axis represents the difference in measurements between the two instruments. The horizontal line centered close to −20 displays the average difference in measurements between the two instruments. The upper and lower, red-dashed lines represent the upper and lower limits, respectively, of the 95% confidence interval for the average difference.

### 3.4 Slide Level Results

Three whole slide images were chosen with a range of UC severity, based on the clinical severity index, pediatric ulcerative colitis activity index (PUCAI) [40]. Specifically, a severe case (PUCAI= 80), a moderate case (PUCAI= 50), and a mild case (PUCAI= 30) were chosen as representative examples. Similar to patch-level annotations, to show the effectiveness of our DL pipeline for eosinophilic cell counts at the whole slide level, we used two trained pathologists’ annotations as comparisons. Note that compared to patch level wherein the counts do not exceed 50 cells, slide-level images can contain hundreds of eosinophil cells and hence require a substantial manual effort, typically an hour for a whole slide image of size 12,000 x 12000 on QuPath/ImageScope. Figure 12 shows the slide-level eosinophilic visualization for the three cases with our DL pipeline-based cell counting against pathologists’ annotations. As can be seen from Figure 12 (Column A) the mild case shows a large presence of eosinophils, Figure 12(Column B) the moderate case shows less eosinophilic density than the mild case, and finally Figure 12 (Column C) the severe case displays significantly less eosinophils that either the mild or moderate cases. These data are consistent with the findings of the PROTECT study [41], that the number of eosinophils in rectal mucosal biopsies decreases with increasing UC severity.

**Figure 12:**
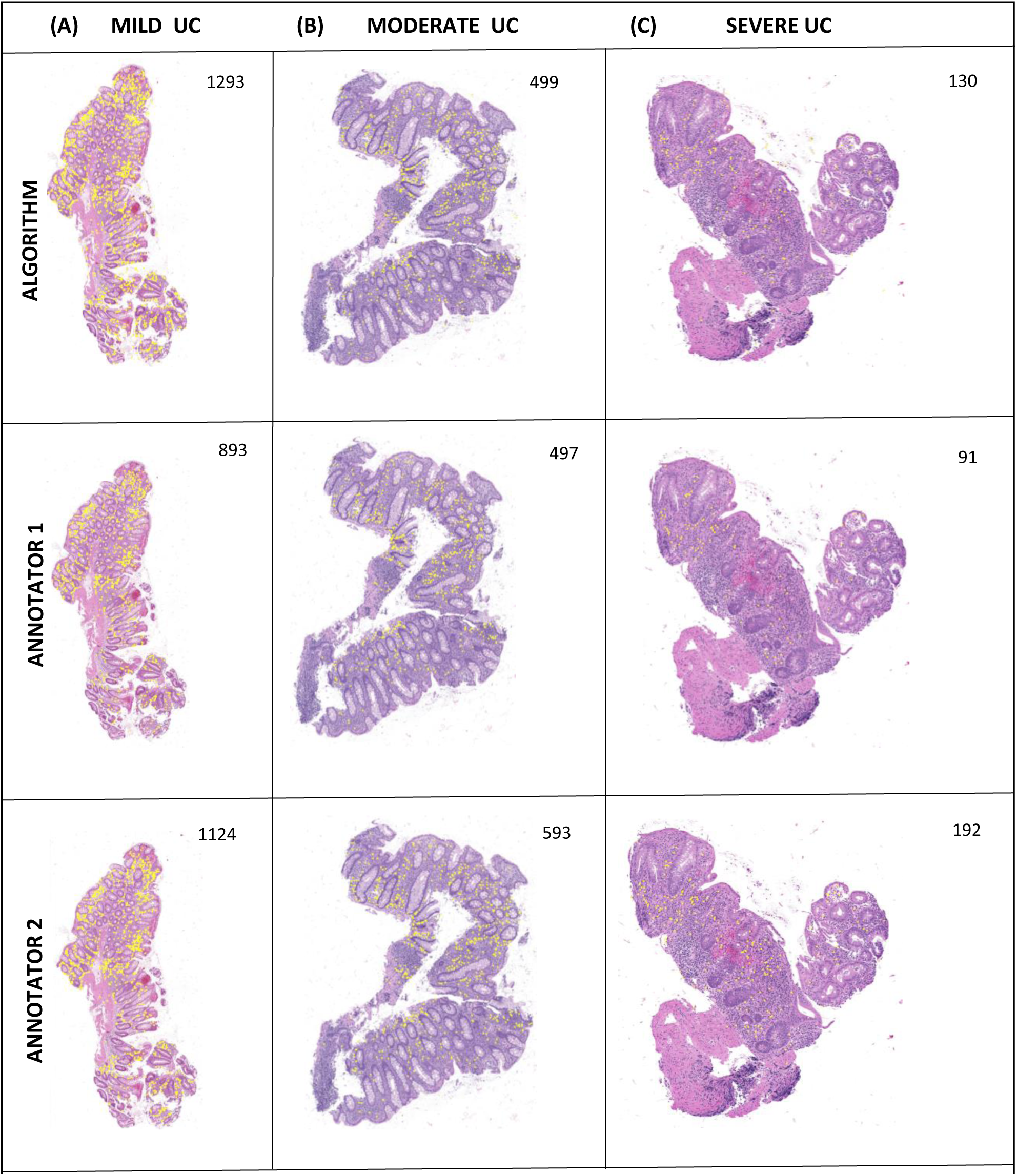
Whole Slide-Level Eosinophilic Visualization for HSK 20x Images (predicted eosinophils shown in yellow). Mild UC (Left), Moderate UC (Middle), and Severe UC (Right). Top row are the algorithmically generated images, while the middle and bottom rows are pathologist-annotated images.

## 4. Discussion and Conclusion

In the present study we have developed a reliable automatic rectal colonic eosinophilic cell counting pipeline. We have utilized a DL approach, extending it to the IBD histology domain. The cell counting pipeline achieved robust and consistent correlation with expert pathologists. Furthermore, the results from our pipeline were comparable, if not superior to the state-of-the-art cell counting algorithm, SAU-Net. Our algorithm achieved an F1-score of 0.86 (95%CI:0.79-0.91) on the test set versus 0.2 (95%CI:0.13-0.26) for SAU-Net. The pipeline demonstrated a high recall of 0.96 (95%CI:0.93-0.99) and a precision of 0.77 (95%CI:0.70-0.83), showing that false positives were more prevalent than false negatives. In addition, the accuracy was similar across the datasets, showing that the model generalized well and did not suffer from overfitting.

It is worth noting that we have developed an initial approach to obtain slide level cell counts by adapting our patch-level model. Whole slide level cell counts were obtained by application of our patch-level model by generating non-overlapping tiles and applying the patch-level pipeline to infer cell counts for each tile. It was not the focus of this study to exhaustively quantify the results of this approach, but qualitatively we have shown that our approach is consistent with the trend of a lower number of eosinophils observed as UC severity increases [41]. It was notable that patch-level training did not limit the accuracy of the final cell counting performance, and in fact, a patch-level analysis is a viable alternative to slide-level approach.

Furthermore, our annotators showed a high degree of consistency as observed by the high Spearman correlation observed for the test set annotations (0.96 (95%CI:0.93-0.97). We are encouraged by this result as it suggests that label noise from inter-annotator variability was limited. As previously mentioned, four experienced pathologists across two institutions were used to provide the ground truth eosinophilic counts for the images. It should be noted that drawing pathologists from the two institutions was done to prevent institutional bias, improve generalizability, and attain inter-laboratory reproducibility. Furthermore, the high degree of correlation among the two pathologists was reflective of their experience in evaluating IBD histopathology. We envisage the automatic cell counting can help standardize eosinophilic cell counting thereby alleviating the subjectivity and variability of traditional manual scoring.

While our pipeline is robust, there are limitations to our approach. First, our work was performed on 20x magnification images. It would be beneficial to extend the work on 40x images as the increased resolution would provide increased granularity; however, 40x histopathology slides are prohibitively large requiring computational power and storage larger than most investigators have available for access. As computing resources develop, this hurdle may be able to be mitigated. A second limitation of our pipeline was that the model was trained on patches instead of the whole slide. A challenge of working with biopsy slides is the sheer size of the digital images. We have overcome both of these related limitations by generating small crops of the whole slide image and training the DL model on these patches as is the standard approach.

We have several extensions to our pipeline that we are developing. First, to validate the pipeline using the dataset from the multicenter inception cohort study, PROTECT [41]. The study was based at 29 centers in the USA and Canada and which included 428 pediatric patients aged 4–17 years with newly diagnosed ulcerative colitis [41]. Following this, the focus will be on whole slide analysis, i.e., computing the eosinophil density of the slide and automating the eosinophilic count per high power field (HPF). The HPF is the field of view at a given magnification the pathologist observes when looking through the viewfinder of the microscope. In cell counting applications, the pathologist scans the slide while viewing through microscope to find the peak region of the slide that exhibits the pertinent phenotype. Work on these extensions is ongoing.

In summary, we have applied a deep learning approach to automate eosinophil counting in UC. Our results suggest that the approach is robust and with further refinement could be applied to other eosinophilic disorders of the gastrointestinal tract, in order to develop a computer aided diagnostic tool in the clinical setting.

## Data Availability

All data produced in the present study are available upon reasonable request to the authors

## Acknowledgement

We thank the Cincinnati Children’s Integrated Pathology Research Facility (Pathology Research Core).

## Author contributions

JR: data curation, analysis and interpretation of data, methodology, software, writing original draft, OLN: acquistion of data, data curation, formal analysis, revision of manuscript ED: design of study, data curation, software, methodology, formal analysis, revision of manuscript DA: data curation and revision of manuscript, XL: data curation, methodology, software, and revision of manuscript. JP: data curation, formal analysis LE: data analysis and interpretation of data, and revision of manuscript. AMG: acquistion of data and revision of manuscript, SP: design of study, supervision and revision of manuscript. IS: acquistion of data, data curation and revision of manuscript JD: conceptualization, design of study, data curation, analysis, funding acquisition, methodology, writing and editing of manuscript and overall study supervision. All authors approved the final version of the manuscript.

## References

1. Hogan, S.P., et al., Eosinophils: biological properties and role in health and disease. Clin Exp Allergy, 2008. 38(5): p. 709–50.

2. Makiyama, K., et al., Activation of eosinophils in the pathophysiology of ulcerative colitis. Journal of gastroenterology, 1995. 30: p. 64–69.

3. Mookhoek, A., et al., The clinical significance of eosinophils in ulcerative colitis: a systematic review. Journal of Crohn’s and Colitis, 2022. 16(8): p. 1321–1334.

4. Ronneberger, O., P. Fischer, and T. Brox. U-net: Convolutional networks for biomedical image segmentation. in Medical Image Computing and Computer-Assisted Intervention– MICCAI 2015: 18th International Conference, Munich, Germany, October 5-9, 2015, Proceedings, Part III 18. 2015. Springer.

5. Komura, D. and S. Ishikawa, Machine learning methods for histopathological image analysis. Computational and structural biotechnology journal, 2018. 16: p. 34–42.

6. Madabhushi, A. and G. Lee, Image analysis and machine learning in digital pathology: Challenges and opportunities. Medical image analysis, 2016. 33: p. 170–175.

7. Stucke, E.M., et al., The value of an additional review for eosinophil quantification in esophageal biopsies. Journal of pediatric gastroenterology and nutrition, 2015. 61(1): p. 65.

8. Xing, F. and L. Yang, Robust nucleus/cell detection and segmentation in digital pathology and microscopy images: a comprehensive review. IEEE reviews in biomedical engineering, 2016. 9: p. 234–263.

9. Maurer, C.R., R. Qi, and V. Raghavan, A linear time algorithm for computing exact Euclidean distance transforms of binary images in arbitrary dimensions. IEEE Transactions on Pattern Analysis and Machine Intelligence, 2003. 25(2): p. 265–270.

10. Soille, P., Morphological image analysis: principles and applications. Vol. 2. 1999: Springer.

11. Raimondo, F., et al., Automated evaluation of Her-2/neu status in breast tissue from fluorescent in situ hybridization images. IEEE Transactions on Image Processing, 2005. 14(9): p. 1288–1299.

12. Byun, J., et al., Automated tool for the detection of cell nuclei in digital microscopic images: application to retinal images. Mol Vis, 2006. 12(105-07): p. 949–60.

13. Matas, J., et al., Robust wide-baseline stereo from maximally stable extremal regions. Image and vision computing, 2004. 22(10): p. 761–767.

14. Duda, R.O. and P.E. Hart, Use of the Hough transformation to detect lines and curves in pictures. Communications of the ACM, 1972. 15(1): p. 11–15.

15. Hough, P.V., Method and means for recognizing complex patterns. 1962, Google Patents.

16. Reisfeld, D., H. Wolfson, and Y. Yeshurun, Context-free attentional operators: The generalized symmetry transform. International Journal of Computer Vision, 1995. 14: p. 119–130.

17. Reisfeld, D. and Y. Yeshurun, Preprocessing of face images: Detection of features and pose normalization. Computer vision and image understanding, 1998. 71(3): p. 413–430.

18. Su, H., et al., Automatic myonuclear detection in isolated single muscle fibers using robust ellipse fitting and sparse representation. IEEE/ACM transactions on computational biology and bioinformatics, 2013. 11(4): p. 714–726.

19. Su, H., et al. Learning based automatic detection of myonuclei in isolated single skeletal muscle fibers using multi-focus image fusion. in 2013 IEEE 10th International Symposium on Biomedical Imaging. 2013. IEEE.

20. SIFT, M.I.U., Automatic Cell Detection in Bright-Field Microscope Images Using SIFT, Random Forests, and Hierarchical Clustering. 2013.

21. Deng, L. and D. Yu, Deep learning: methods and applications. Foundations and trends® in signal processing, 2014. 7(3–4): p. 197–387.

22. LeCun, Y., Y. Bengio, and G. Hinton, Deep learning. nature, 2015. 521(7553): p. 436–444.

23. Roth, H.R., et al. A new 2.5 D representation for lymph node detection using random sets of deep convolutional neural network observations. in Medical Image Computing and Computer-Assisted Intervention–MICCAI 2014: 17th International Conference, Boston, MA, USA, September 14-18, 2014, Proceedings, Part I 17. 2014. Springer.

24. Shen, D., G. Wu, and H.-I. Suk, Deep learning in medical image analysis. Annual review of biomedical engineering, 2017. 19: p. 221–248.

25. Wang, R., et al., Medical image segmentation using deep learning: A survey. IET Image Processing, 2022. 16(5): p. 1243–1267.

26. Meijering, E., Cell segmentation: 50 years down the road [life sciences]. IEEE signal processing magazine, 2012. 29(5): p. 140–145.

27. Falk, T., et al., U-Net: deep learning for cell counting, detection, and morphometry. Nature methods, 2019. 16(1): p. 67–70.

28. Srinidhi, C.L., O. Ciga, and A.L. Martel, Deep neural network models for computational histopathology: A survey. Medical Image Analysis, 2021. 67: p. 101813.

29. Guo, Y., et al., SAU-net: a unified network for cell counting in 2D and 3D microscopy images. IEEE/ACM transactions on computational biology and bioinformatics, 2021. 19(4): p. 1920–1932.

30. Paszke, A., et al., Pytorch: An imperative style, high-performance deep learning library. Advances in neural information processing systems, 2019. 32.

31. Feldman, A.T. and D. Wolfe, Tissue processing and hematoxylin and eosin staining. Histopathology: Methods and Protocols, 2014: p. 31–43.

32. Salvi, M., et al., The impact of pre-and post-image processing techniques on deep learning frameworks: A comprehensive review for digital pathology image analysis. Computers in Biology and Medicine, 2021. 128: p. 104129.

33. Bradski, G., The openCV library. Dr. Dobb’s Journal: Software Tools for the Professional Programmer, 2000. 25(11): p. 120–123.

34. Harris, C.R., et al., Array programming with NumPy. Nature, 2020. 585(7825): p. 357–362.

35. Bankhead, P., et al., QuPath: Open source software for digital pathology image analysis. Scientific reports, 2017. 7(1): p. 1–7.

36. Adorno III, W., et al. Advancing eosinophilic esophagitis diagnosis and phenotype assessment with deep learning computer vision. in Biomedical engineering systems and technologies, international joint conference, BIOSTEC… revised selected papers. BIOSTEC (Conference). 2021. NIH Public Access.

37. Chang, H.Y., et al., Artificial intelligence in pathology. Journal of pathology and translational medicine, 2019. 53(1): p. 1–12.

38. Lu, M.Y., et al., Data-efficient and weakly supervised computational pathology on whole-slide images. Nature biomedical engineering, 2021. 5(6): p. 555–570.

39. Van der Laak, J., G. Litjens, and F. Ciompi, Deep learning in histopathology: the path to the clinic. Nature medicine, 2021. 27(5): p. 775–784.

40. Turner, D., Severe acute ulcerative colitis: the pediatric perspective. Digestive Diseases, 2009. 27(3): p. 322–326.

41. Hyams, J.S., et al., Factors associated with early outcomes following standardised therapy in children with ulcerative colitis (PROTECT): a multicentre inception cohort study. The Lancet Gastroenterology & Hepatology, 2017. 2(12): p. 855–868.

